# Development and Validation of a Machine Learning Algorithm to Classify Lower Urinary Tract Symptoms

**DOI:** 10.1101/2022.12.25.22283168

**Authors:** Jeffrey N. Chiang, Kai B. Dallas, Ashley T. Caron, Jennifer T. Anger, Melissa R. Kaufman, A. Lenore Ackerman

**Author notes:** Corresponding Author: A. Lenore Ackerman, MD, PhD, Director of Research, Division of Pelvic Medicine and Reconstructive Surgery, Assistant Professor of Urology and Obstetrics and Gynecology, David Geffen School of Medicine at UCLA, Box 951738 • Los Angeles, California • 90095-1738.

## Abstract

**Objective:** Lower urinary tract symptoms (LUTS), such as urinary urgency, frequency, and incontinence, affect the majority of the population, causing substantial morbidity, yet few receive effective care. Sizeable symptomatic overlap between LUTS categories leads to high rates of misdiagnosis. To improve diagnostic accuracy, we sought to employ machine learning approaches to LUTS categorization to generate diagnostic groupings based on patient-reported clinical data, creating a novel tool for diagnosis of patients with voiding complaints.

**Methods:** Questionnaire responses in a Development Dataset of 514 female subjects were used for model development, identifying 4 major clusters and 9 specific phenotypes of LUTS using agglomerative hierarchical clustering. Each cluster and phenotype was assigned a clinical identity consistent with recognized causes of voiding dysfunction by the consensus of two urologic specialists. Then, a random forest classifier was trained to assign unseen patients into these phenotypes. That model was then applied to a Validation Dataset of 571 additional subjects to confirm the diagnostic algorithm.

**Results:** This data-driven, hierarchical clustering approach captured overlapping symptoms inherent in typical patients, recognizing common uncomplicated diagnoses (i.e., overactive bladder) as well as several underrecognized diagnostic categories (i.e., myofascial pelvic pain). A diagnostic algorithm derived by supervised machine learning to assign unseen subjects into these phenotypes demonstrated good reproducibillty of the phenotypes and their symptomatic patterns in an independent Validation Dataset.

**Conclusions:** We describe the generation of a machine learning algorithm relying only on validated, patient-reported symptoms for diagnostic classification. Given a growing physician shortage and increasing challenges for patients accessing specialist care, this type of digital technology holds great potential to improve the recognition and diagnosis of functional urologic conditions.

## Introduction

Half of adult women are incontinent; more than 75% of women at some point in the lifespan report disruptions in daily activities by at least one lower urinary tract symptom (LUTS), such as *urinary urgency, frequency, nocturia, painful urination, bladder pain or discomfort, or incontinence*.^1,2^ When severe, these urinary symptoms degrade health-related quality of life to levels worse than chronic dialysis.^3^ Incontinence and urinary complaints are one of the most common causes for the long-term institutionalization of older adults.^4^ Further, these disorders represent substantial economic burden, with annual costs estimated to approach 100 billion dollars.^5^ Yet despite this large burden of illness, most women do not seek care, due to embarrassment, resignation, or misconceptions that these symptoms are normal or cannot be treated.^6^

Even for those who do seek care, many will not receive an accurate diagnosis.^7^ While patients presenting with urinary symptoms are categorized into several unique conditions, appropriate diagnosis and treatment is complicated by subjectivity of language used to describe symptoms and sizeable symptomatic overlap.^8^ While classified as separate conditions, diagnoses such as overactive bladder (OAB) and interstitial cystitis/painful bladder syndrome (IC/BPS) are not diseases, but symptom complexes without a known pathophysiology that share many overlapping symptoms. Clinicians typically assign a diagnosis according to patients’ most bothersome complaints, but as there are currently no definitive tests or biomarkers available, diagnosis and treatment assignment are subjective, based entirely on clinician judgement.^9,10^ Due to poor efficacy and treatment side effects, more than 90% of patients will abandon medical therapies within a year.^11^

A data-driven approach may address many of the current obstacles to LUTS care. Virtual screening approaches would allow anonymized recognition of symptoms without requiring in-person or specialist visits. Given the high degree of overlapping symptoms, but distinctive patterns of combined symptoms, algorithms identifying more specific phenotypes of LUTS may recognize subtle distinctions indicative of unique causes of convergent symptomatologies that clinicians may have difficulty recognizing. While symptoms of LUTS diagnoses may be similar, the underlying etiologies of these symptoms may be quite different and require different approaches to treatment. To this end, multiple studies have attempted to utilize a range of unsupervised machine learning (ML) methods to improve phenotypic classification of lower urinary tract disorders. However, most have examined only a subset of symptoms, typically excluding patients with complex symptoms or confounding factors such as pelvic organ prolapse (POP). However, “as often patients present with multiple urinary symptoms that do not perfectly fit the pre-established diagnoses,”^12^ the intentional exclusion in these studies of patients with complex or overlapping symptoms or co-existing pelvic organ prolapse may underlie the lack of practical progress in improving current diagnostic schema.

For most ML methods, data objects are divided into non-overlapping, mutually-exclusive clusters of a fixed number. In contrast, hierarchical clustering generates a set of nested clusters that are arranged as a tree/dendrogram. Hierarchical clustering is best used when the number of classes is not known; once the structure of the data is defined, one can stop at any number of clusters. A strength of hierarchical clustering is the ability for the resulting dendrogram to provide insights as to the structure of the data, which in turn informs about the number of logical clusters. Given the nature of functional urologic symptoms, we believe that hierarchical clustering presents a more complete picture of the underlying patterns of lower urinary tract symptoms than other methods of machine learning. In addition, It may provide a more useful framework to determine what level of phenotypic classification is useful in clinical practice.

Given the known obstacles and clinical dilemmas in diagnosing LUTS, we aimed to create an unsupervised ML algorithm encompassing all patients presenting with LUTS. This novel tool for the diagnosis of patients with voiding complaints utilized agglomerative hierarchical clustering to generate diagnostic groupings based on patient-reported clinical data.

## Methods

### Study Cohorts

After Institutional Review Board (IRB#00040261) approval, the *Development Dataset* of 514 female subjects with a wide range of symptoms, seen consecutively for evaluation in a tertiary urogynecology practice between January and December 2018, was used for model development. A second *Validation Dataset* of 571 subjects recruited from the same practice consequtively between January 2019 and March 2020 was used to validate the diagnostic ML algorithm. For each cohort, subjects were administered four, validated questionnaires at initial evaluation: the female Genitourinary Pain Index (fGUPI)^13^, Overactive Bladder Questionnaire (OABq)^14^, Pelvic Floor Distress Inventory (PFDI-20)^15^ and O’Leary-Sant Indices, including the Interstitial Cystitis Symptom and Problem Indices (ICSI/ICPI).^16^ The fGUPI measures the nature and severity of genitourinary pain, and contains subscales assessing pain, urinary symptoms, and quality of life.^13^ The OABq measures continent and incontinent OAB symptoms, symptom bother, coping behaviors, concern/worry, social interaction, sleep, and health-related quality of life.^14^ Only the symptom-based questions (1-8) were utilized in this analysis. The PFDI-20 measures urinary, defecatory, and prolapse symptoms commonly associated with pelvic floor disorders such as POP.^15^ The ICSI/ICPI are used together to measure the severity and bother of urinary frequency, urgency, nocturia, and bladder pain.^16^ Patients with active urinary tract infection (UTI), prior pelvic reconstructive surgeries (such as prolapse repairs, bladder augmentation, incontinence procedures), current pregnancy, or neurogenic bladder were excluded from the study. Patients with cyclic pain at menses were excluded, however, patients with prior history of UTI or comorbid functional pain syndromes, such as irritable bowel syndrome or fibromyalgia, were allowed to participate. Patients with asymptomatic complaints (e.g., microscopic hematuria) were included in both datasets.

### Hierarchical Clustering

Agglomerative hierarchical clustering was applied to the *Development Dataset* to identify patient groups on the basis of patient age and questionnaire responses. This unsupervised approach recursively builds a hierarchical representation from the “bottom up” by grouping pairs of samples according to a distance metric and linkage criterion. This algorithm was applied using the Ward linkage criterion, which aims to create groupings which have minimum intra-group variance.^17^ Similarity between samples was calculated using the Euclidean (i.e., squared) distance between symptoms. After the hierarchical representation was constructed using the agglomerative algorithm, clusters were created by truncating the depth of the hierarchical representation, thereby grouping patients into a fixed number of clusters (e.g., **Fig. 1**). Mean silhouette scores^18^ were computed to measure cluster assignment performance to select the optimal cluster assignment between 2 and 15 clusters. Using this criterion, four and nine cluster solutions were selected.

**Figure 1.**
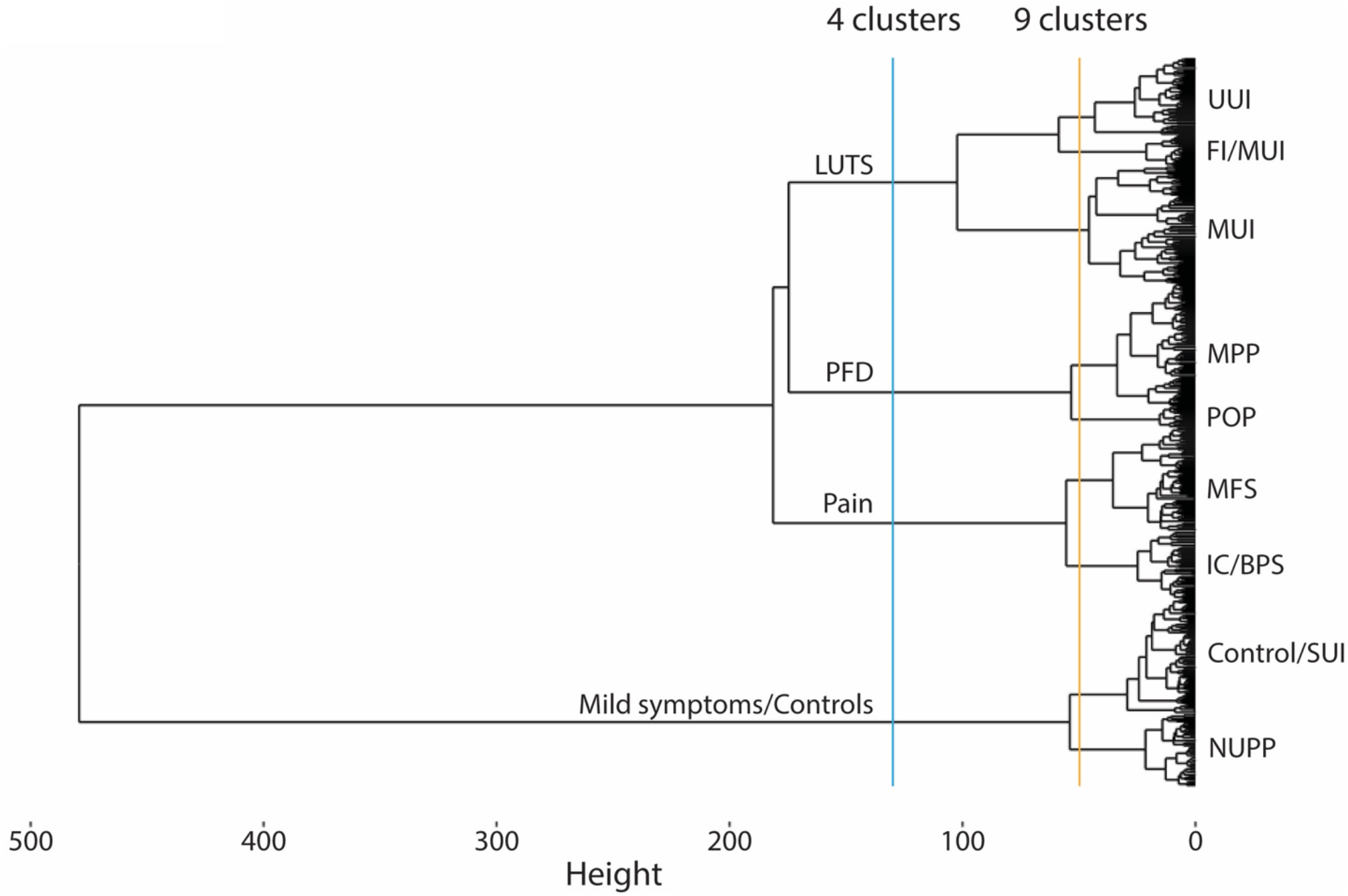
Dendrogram indicating the hierarchical relationships between clusters. The different lines indicate different possible cut-off points used to subclassify subjects with LUTS, with the blue line indicating the major symptomatic divisions in subjects presenting for care in a urogynecology clinic (*LUTS*: lower urinary tract symptoms; *PFD*: pelvic floor disorders; *Pain*: pelvic pain complaints; *Mild symptoms/controls*: patients with mild or minimal complaints). The yellow line indiciates the location in the dendrogram at which subjects are separated into 9 unique clusters, with each of the major categories further divided into 2-3 subclusters (UUI: urgency urinary incontinence; MUI: mixed urinary incontinence; FI/MUI: fecal incontinence/UUI; MFS: myofascial frequency syndrome; POP: pelvic organ prolapse; MPP: myofascial pelvic pain; IC/BPS: interstitial cystitis/bladder pain syndrome; NUPP: non-urologic pelvic pain; Control/SUI: Asymptomatic subject/stress urinary incontinence).

### Phenotypic group descriptive assignments

Age and survey response scores were scaled to a set range before plotting as a heat map. ANOVA assessed significant differences between cluster groups in intragroup means for each variable. The dominant features for each cluster and phenotype were examined independently by two urogynecologists, who each assigned a clinical identity based on their expert opinion. For any disagreement, discussion between the two evaluators and a third moderating urogynecologist allowed for refinement of the group clinical identity until consensus was reached.

### Generation of the diagnostic algorithm

Unsupervised algorithms, such as agglomerative clustering, are not reliably applicable to new data as they require the entire hierarchical representation to be re-generated each time the algorithm is run. After manually validating cluster assignments and assigning phenotypic group descriptions, a supervised machine learning model was trained to assign unseen patients into existing clusters. Using the Development Dataset, random forest models were trained to predict the machine-generated cluster assignment on the same symptoms used for clustering. Random forests are generally robust to overfitting and outliers, and outperform other methods when representing tabular data.^19^ Two models were generated which assigned patients into the four and nine cluster labels, respectively. These models were assessed using the validation dataset with balanced accuracy^20^ (defined as the average sensitivity score for each class) – which has been used to deal with imbalanced data.

### Uniform manifold approximation and projection (UMAP) visualization

UMAP is a nonlinear dimension reduction technique used to visualize high-dimensionality data for qualitative inspection.^21^ Using the development dataset, the algorithm was applied to represent patients in two data-generated axes. As in the clustering analysis, the projection axes were optimized with respect to the Euclidean distance between patient responses. Then, Validation Dataset patients were transformed into the same space and all patients visualized together. Each patient was colored according to the phenotype assigned by the nine-cluster model.

### Statistical analysis

Cluster fitting and optimization, machine learning model development and evaluation, and UMAP were performed with Python using the scikit-learn and UMAP-learn packages. All other analyses and visualization were performed in R version 4.2.1. Differences in demographic and clinical characteristics were compared by using Wilcoxon signed rank tests for paired data and the Pearson chi square, Fisher exact, or Mann-Whitney U-tests for independent data as appropriate (2-tailed). Differences in proportions were compared using the two-sample z-test. Results were considered significant at an alpha level <0.05.

## Results

### Unsupervised clustering of subjects presenting with LUTS

The *Development Dataset* of 514 female patients with a mean age of 58.7 years completed a panel of validated symptomatic questionnaires assessing genitourinary symptoms between June 2017 and December 2018. Using only age and patient-reported symptoms, subjects were classified into symptomatic clusters according to an agglomerative hierarchical clustering algorithm (**Fig. 1**). Examination of the resulting dendrogram (**Fig. 1**, blue line) revealed four clear branches, confirmed as the optimal number of clusters using the silhouette method (global maximum, **Fig. 2**).

**Figure 2:**
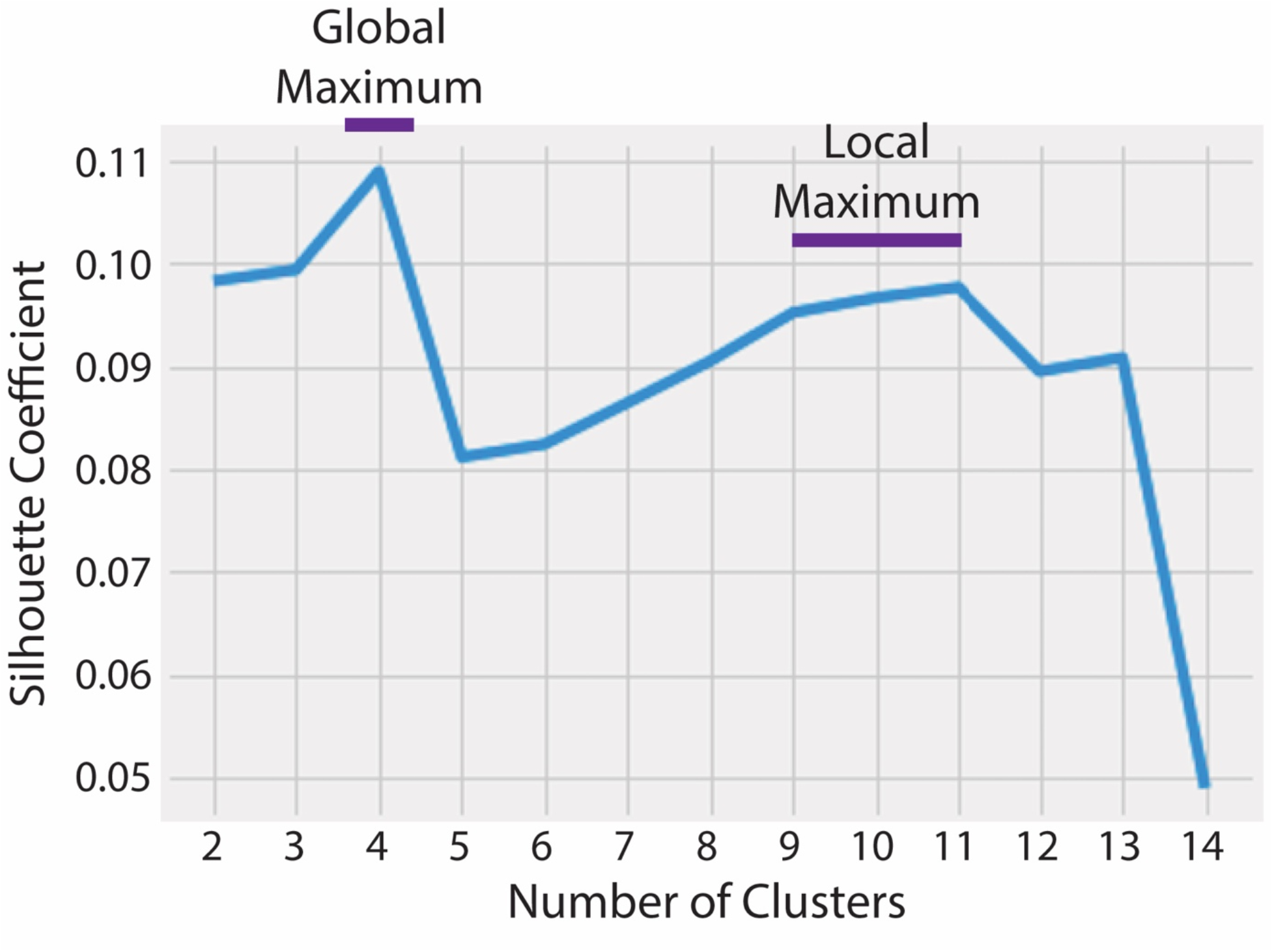
Optimal number of clusters for the hierarchical clustering by the silhouette method. The Silhouette score serves as a measure of the goodness of a clustering technique, encompassing both how similar each individual is to others in the same cluster (cohesion) compared to others in other distinct clusters (separation). Higher value scores indicate that each individual is well matched to their own cluster and poorly matched to neighboring clusters.

### Data-driven hierarchical clustering reveals the global patterns in phenotypes

The symptom patterns associated with each cluster (**Table 1**) reflected the overall categories of subjects seeking urogynecologic care: 1) subjects with mild symptoms who are only moderately bothered (*Mild symptoms/controls*), 2) subjects with predominantly lower urinary tract symptoms, such as urinary frequency, urgency, and incontinence (*LUTS*), 3) subjects with pain as their predominant complaint (*Pain*), and 4) subjects with pelvic floor complaints (pelvic pressure, vaginal bulge) who also exhibit urinary symptoms and discomfort (pelvic floor disorders [*PFD*]). These patterns are more clearly visualized in a heat map (**Fig. 3**) demonstrating which features are prominent in each group. The *PFD* group had globally elevated symptom scores across all symptomatic domains, with unique complaints attributable to the pelvic floor, such as defecatory dysfunction (PFDI20 q7-8), vaginal bulge (PFDI20 q3), pelvic pressure (PFDI20 q2), and a need to splint to defecate or void (PFDI20 q4, PFDI20 q6). The *Pain* group was the youngest group overall and displayed similarly high scores to the *PFD* group only on questions assessing pelvic and genitourinary pain (fGUPI q1-4, PFDI20 q20, ICSI q4, ICPI q4). The *LUTS* group was generally older than the other groups and exhibited elevated urinary incontinence related to both urgency (PFDI20 q16, OABq q8) and stress (activities that increase intraabdominal pressure) (OABq q4, PFDI20 q17). While these subjects commonly exhibited urinary urgency (OABq q3, ICPI q3, ICSI q1), frequency (ICSI q2, ICPI q1, GUPI q6), and nocturia (ICPI q2, OABq q5) at levels greater than the controls, symptom severities of the *LUTS* group only surpassed the *PFD* group in the domain of fecal incontinence (PFDI20 q9-11).

**Table 1.**
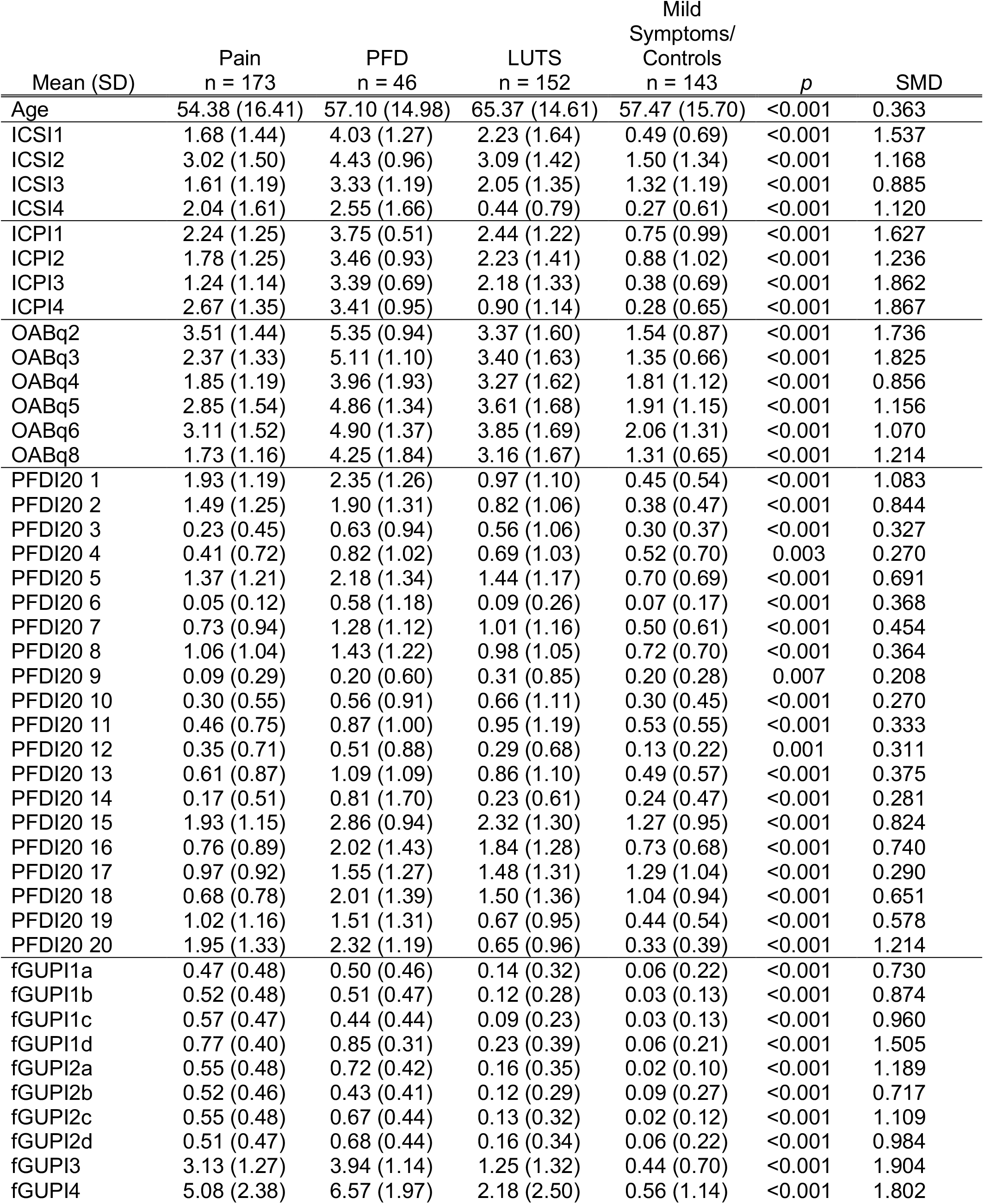
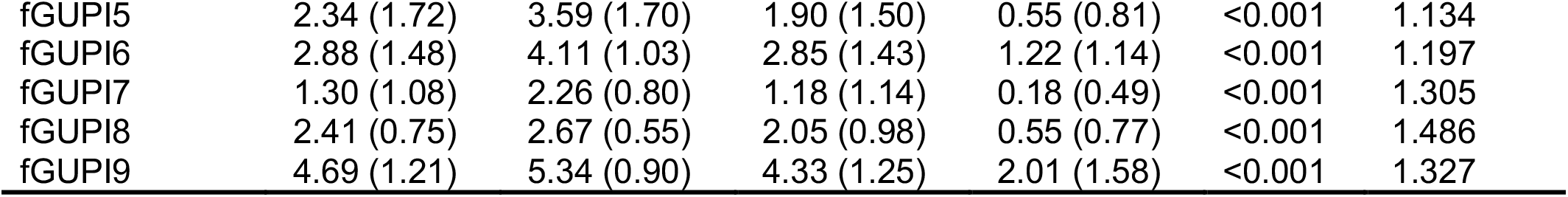
*Patient survey scores for subjects seeking care in a urogynecology clinic by* 4-cluster machine learning diagnostic groups. Means and standard deviations are reported for each group. *LUTS*: lower urinary tract symptoms; *PFD*: pelvic floor disorders; *Pain*: pelvic pain complaints; *Mild symptoms/controls*: patients with mild or minimal complaints. *SMD*: standard mean difference.

**Figure 3.**
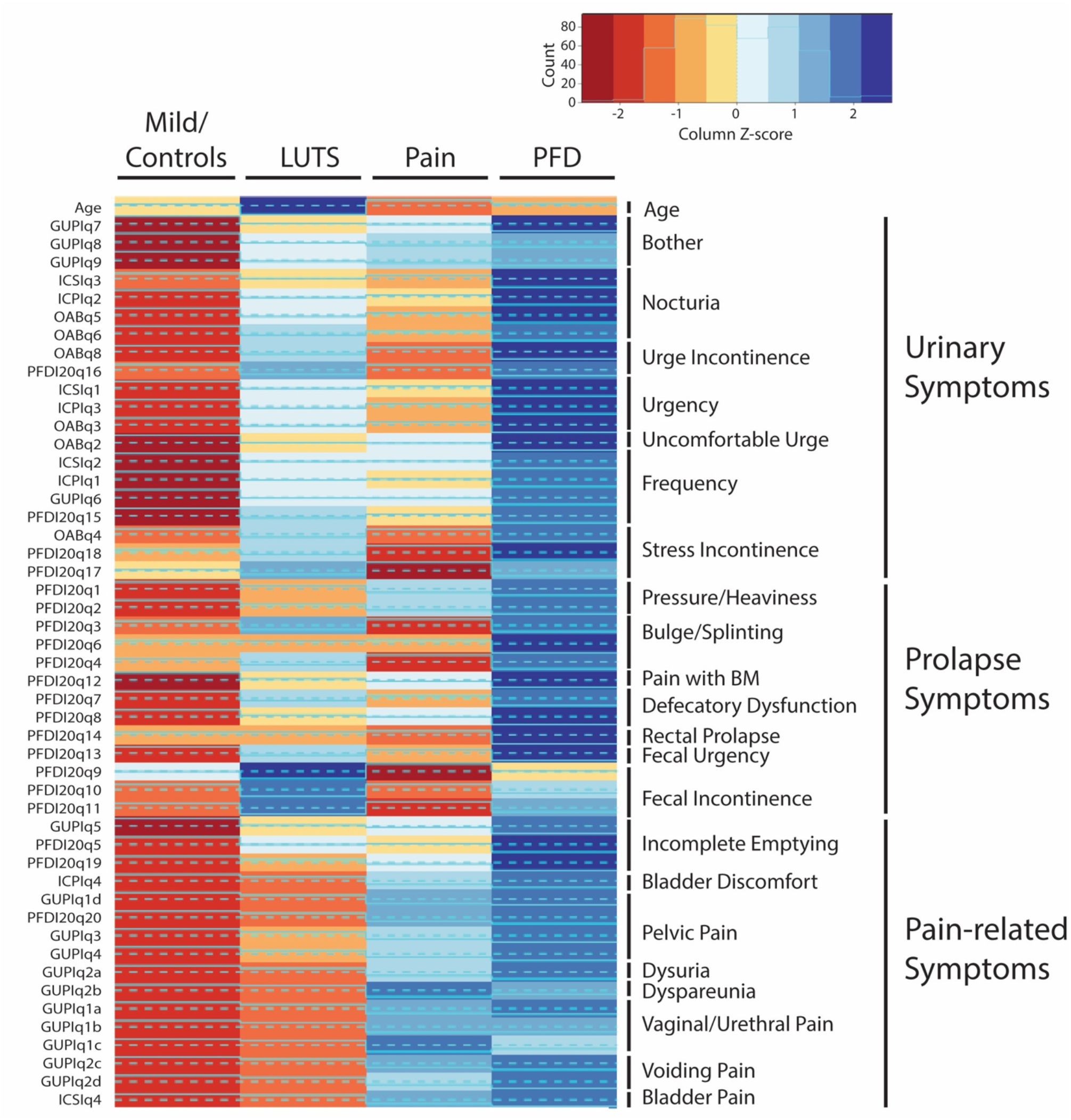
Heatmap of survey responses and patient characteristics for each of the machine learning generated clusters for the 4 major clusters. Specific symptomatic patterns for the 4 clusters (blue in Figure A), with deepening blues indicating scores increasingly elevated over the population mean and deeper shades of red indicating scores increasingly lower than the population mean. (*LUTS*: lower urinary tract symptoms; *PFD*: pelvic floor disorders; *Pain*: pelvic pain complaints; *Mild symptoms/controls*: patients with mild or minimal complaints).

### Discrimination of symptomatic phenotypes

While the four-cluster classification provides a general categorization of women presenting for urogynecologic care, the resulting groups encapsulate only the most basic symptomatic categorization, failing to distinguish patients for whom different treatments are typically selected, such as stress and urgency incontinence, vestibulodynia and interstitial cystitis/bladder pain syndrome, or pelvic organ prolapse and myofascial pelvic pain. We therefore examined the next most optimal range of clusters. The silhouette score reached a local maximum in the range of 9-11 clusters (**Fig. 2**, local maximum); in the dendrogram, the nine-cluster division best captured the major subdivisions in the data (**Fig. 1**, gold line). Again visualized as a heat map, patterns of dominant symptoms provide characterization of each cluster phenotype (**Table 2**), each of which was assigned a phenotypic designation by specialist consensus (**Fig. 4**).

**Table 2.**
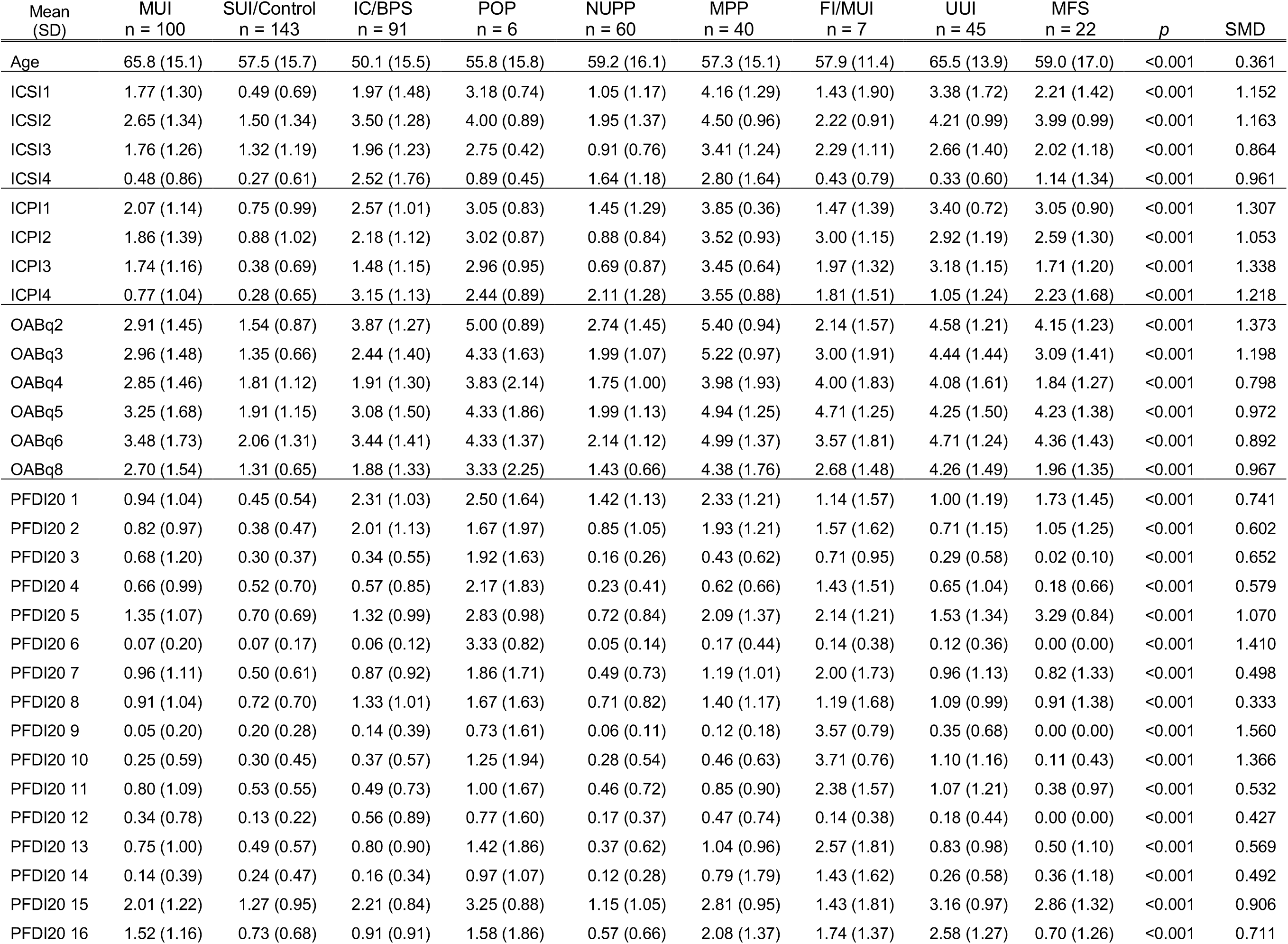
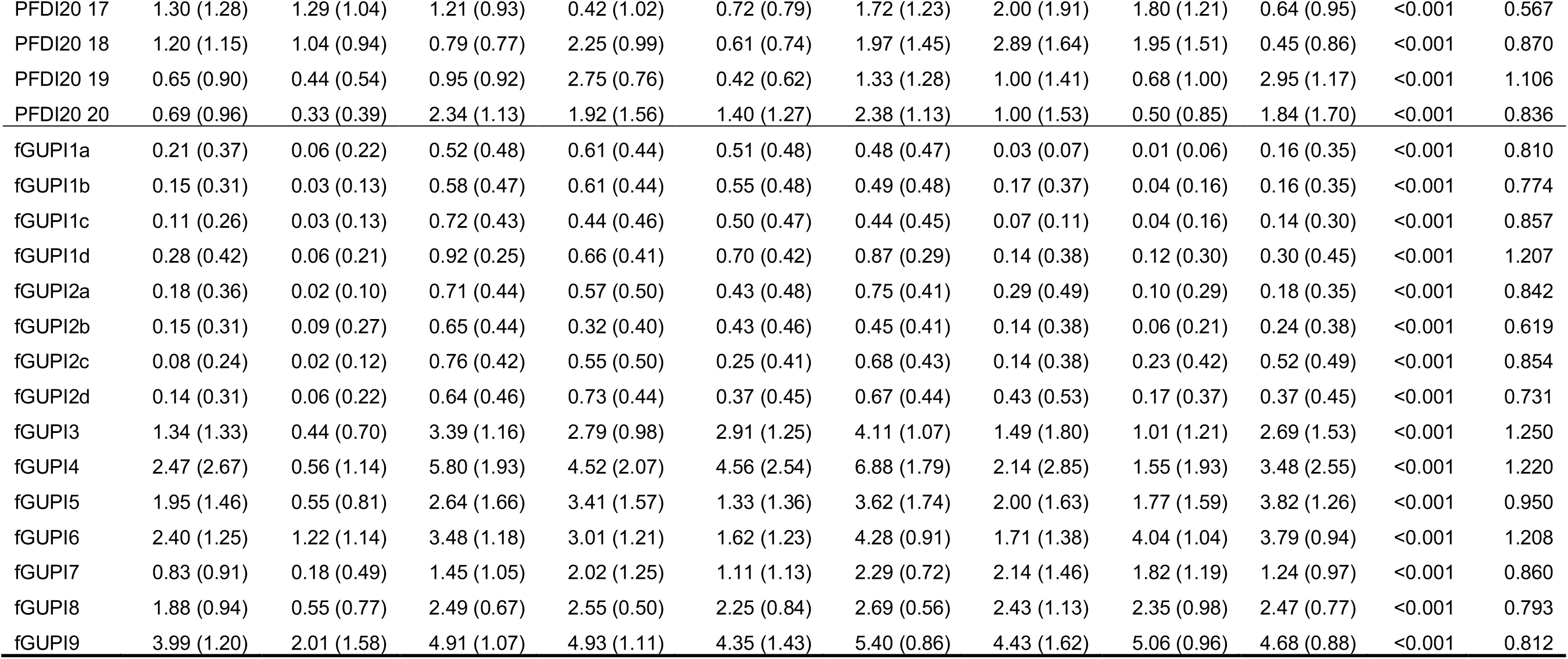
Group symptom means and differences for 9 symptom subclusters of subjects seeking care in a urogynecology clinic. Groups means and standard deviations are reported for each group. MUI: mixes urinary incontinence; Control/SUI: Asymptomatic subject/stress urinary incontinence; IC/BPS: interstitial cystitis/bladder pain syndrome; POP: pelvic organ prolapse; NUPP: non-urologic pelvic pain; MPP: myofascial pelvic pain; FI/MUI: fecal incontinence/mixed urinary incontinence; UUI: urgency urinary incontinence; MFS: myofascial frequency syndrome. SMD: Standard mean difference.

**Figure 4:**
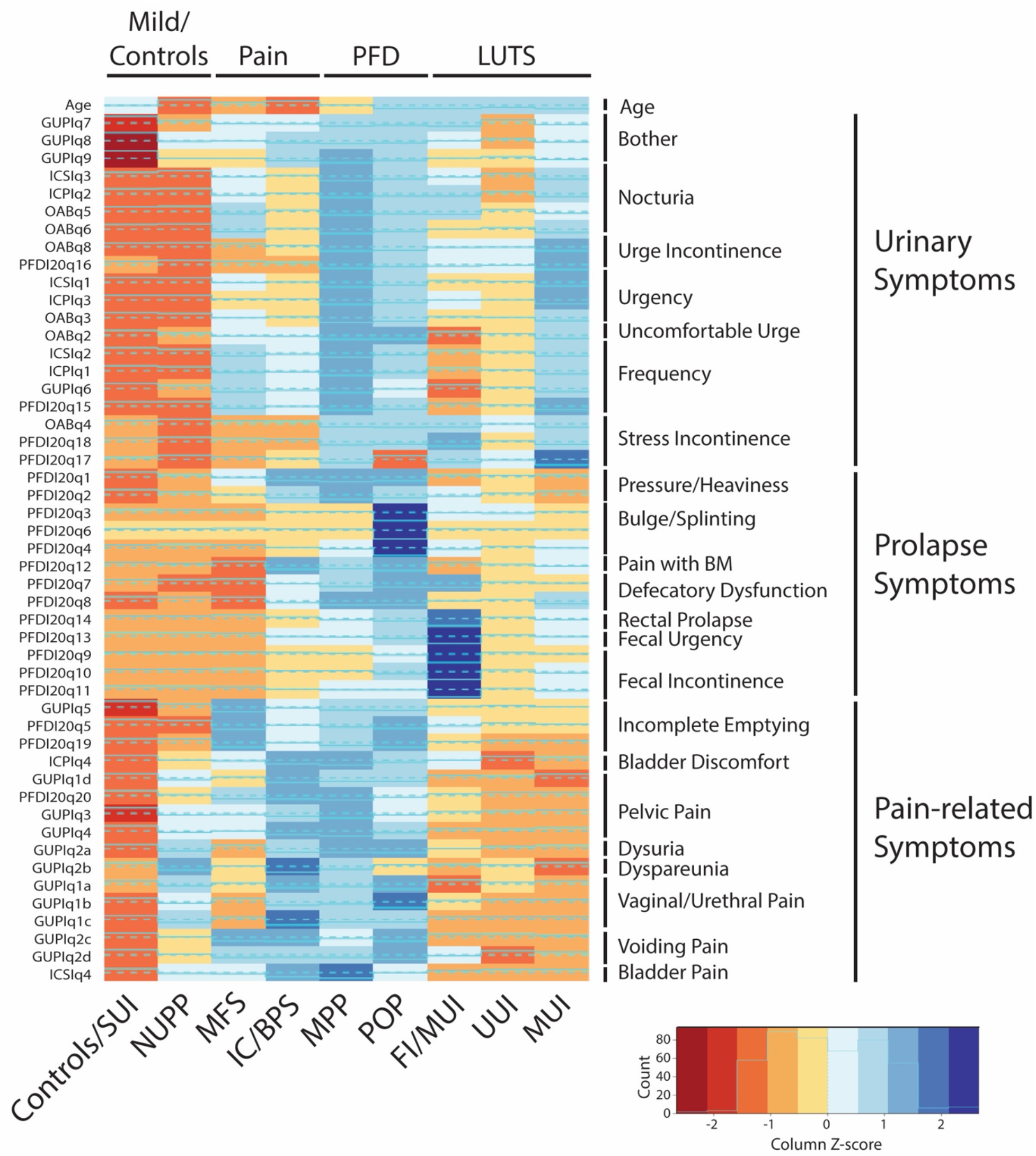
Heatmap of survey responses and patient characteristics for each of the machine learning generated clusters. Specific symptomatic patterns for the nine clusters shown in Figure A, with deepening blues indicating scores increasingly elevated over the population mean and deeper shades of red indicating scores increasingly lower than the population mean. (UUI: urgency urinary incontinence; FI/UUI: fecal incontinence/UUI; MUI: mixes urinary incontinence;’ HT PFD: high-tone pelvic floor dysfuction; POP: pelvic organ prolapse; MPP: myofascial pelvic pain; IC/BPS: interstitial cystitis/bladder pain syndrome; NUPP: non-urologic pelvic pain; Control/SUI: Asymptomatic subject/stress urinary incontinence).

The *LUTS* cluster was subdivided into three phenotypes: 1) urgency urinary incontinence (*UUI*), who lacked other bothersome sypmtoms, 2) mixed urinary incontinence (*MUI*), who exhibited both UUI and SUI, and 3) fecal incontinence/mixed urinary incontinence (*FI/MUI*), who were profoundly impacted by both fecal urgency and incontinence in addition to their urinary symptoms. The *Pain* cluster was subdivided into an *IC/BPS* phenotype, who were bothered by classic bladder pain with bladder filling, relieved by bladder emptying, and a second phenotype whose symptoms were described more as bladder pressure and discomfort. The co-existence of these symptoms with a strong sensation of incomplete bladder emptying identified this group as myofascial frequency syndrome (*MFS*), a condition in which urinary symptoms are caused by pelvic floor myofascial dysfunction. The *PFD* cluster was further divided into subjects with clear pelvic organ prolapse (*POP*), who described both a bothersome vaginal bulge and needing to reduce the prolapse to urinate or defecate, and a myofascial pelvic pain (*MPP*) group, who have both genitourinary pain and global urinary, defecatory, and sexual dysfunction, similar to the POP group, but who lack any evidence of vaginal bulge. The last cluster (*Mild symptoms/controls*) included two phenotypes of subjects: those with stress urinary incontinence (*Controls/SUI*) who were often only minimally bothered, and those with focal genital pain unrelated to bladder filling or voiding, known as non-urologic pelvic pain (*NUPP*).

### Development of a classification algorithm

We next sought to create a diagnostic tool for the phenotypic classification of new patients. We trained a random forest classifier using the phenotypic assignments of the Development Dataset to create a classification algorithm, which we then applied to the independent Validation Dataset of 571 individuals with urologic complaints. Phenotypic assignment using the random forest classifier resulted in groups that were highly similar to the development dataset phenotypes, exhibiting the same patterns of distinctive symptoms. We plotted the similarity of the development and validation phenotypes graphically using a uniform manifold approximation and projection (UMAP) method (**Fig. 5**). Proximity in UMAP space signifies similarity between groups, in this case capturing the overlap in symptom profiles between phenotypes from the validation and development cohorts as well as the relationships between groups. For example, the mixed urinary incontinence (MUI) group, which exhibits incontinence elicited by urgency and stress, exists at the interface between the urgency urinary incontinence (UUI) and stress urinary incontinence (SUI) groups, sharing features of both while still occupying a unique space.

**Figure 5:**
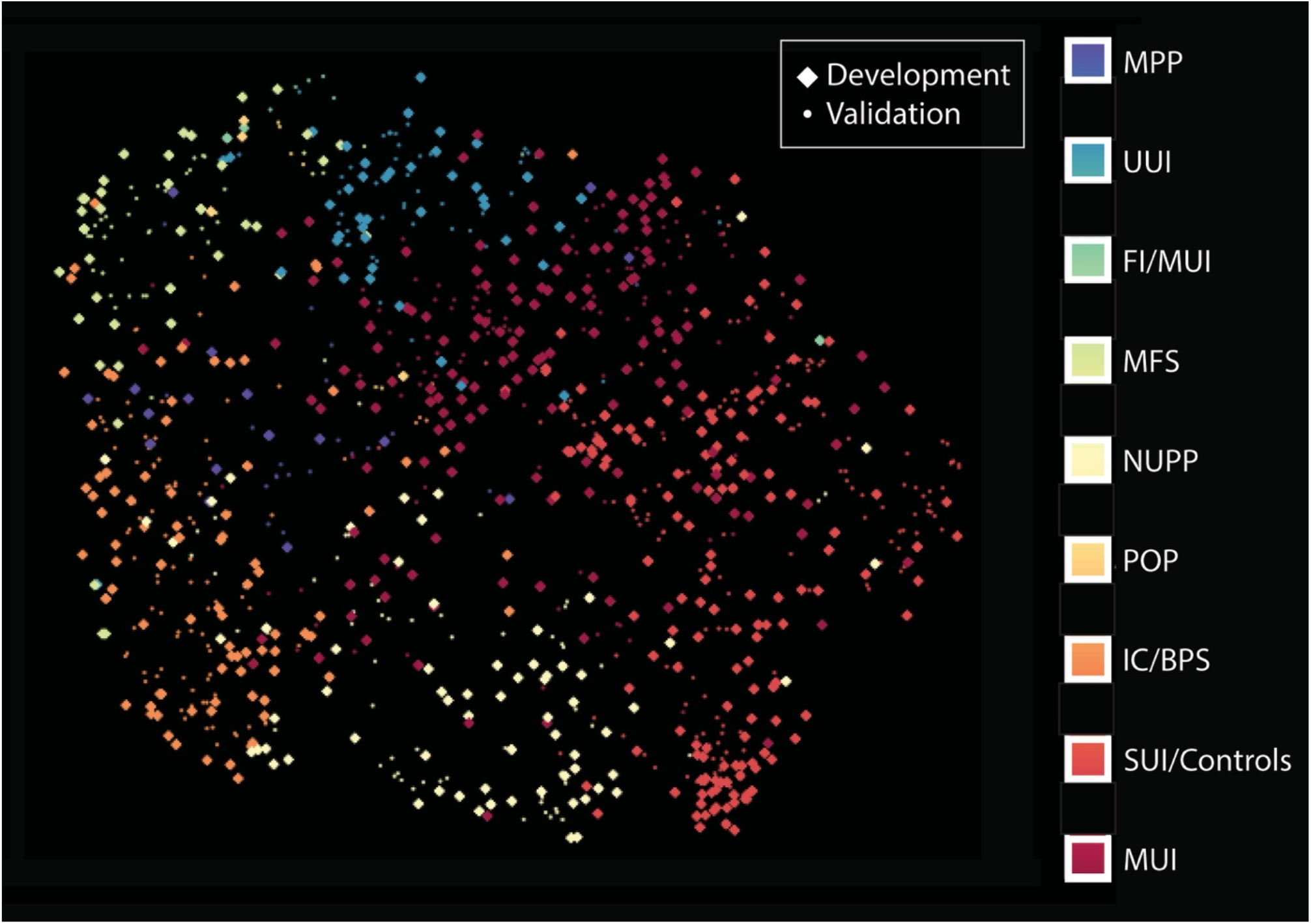
Uniform Manifold Approximation and Projection (UMAP) plot visualizing the overlap in clusters with respect to the symptomatic features between the development and validation populations. Good overlap was seen between the clusters in the development (larger diamonds) and validation (smaller circles) cohorts.

## Discussion

We successfully applied machine learning algorithms to the diagnostic classification of women with a wide range of symptoms presenting for urologic care. This classification generated logical, phenotypic groups based on validated, patient-reported symptoms alone. Symptomatic patterns could be grouped into four general clusters: minimal/mild symptoms, urogenital pain, urinary complaints, and pelvic floor disorders. Validation of these clusters revealed high reproducibility in an independent cohort. These groups are analogous to the general clinical categories currently used; most patients presenting for urologic care will be diagnosed with either incontinence, genitourinary pain, or pelvic organ prolapse. As unsupervised machine learning brings no assumptions to cluster derivation, agreement of the overall diagnostic categories with well-accepted clinical categorization validates the ability of data-driven methods to derive clinically meaningful diagnostic categories.

Several groups have tried to subclassify urologic phenotypes, but have typically examined only one symptom cluster in isolation (e.g., overactive bladder^12,22-24^ or genitourinary pain^25-28^). Substantial work, however, documents that these divisions are artificial; many patients carrying an OAB diagnosis suffer from significant bladder pain (35%) while more than 30% of IC/BPS patients have urge incontinence.^29,30^ In addition, OAB symptoms are common in patients desiring surgical repairs for POP.^31^ Given the high prevalence of prolapse in the population (24-49%),^32-34^ it is unclear how co-existing POP should influence our clinical management of patients presenting primarily with urinary complaints.

To overcome this obstacle, broad inclusion of all patients consecutively presenting for urogynecologic care combined with unsupervised clustering using patient complaints alone allowed us to derive nine unique phenotypes encompassing the range of overlapping symptoms without bias. Distinction between groups was based on unique combinations of symptoms rather than individual, pathognomonic features.^23^ These nine phenotypic diagnoses included the range of common urologic diagnoses (SUI, UUI, MUI, IC/BPS), but also incorporated several less common, emerging pathologies that have only recently been suspected in the etiology of LUTS (MFS, MPP, NUPP).^28,35-37^ The classifier also distinguished between subjects with mixed urinary incontinence in whom a correctable, anatomic cause (POP) to their symptoms should be suspected, which may influence treatment choices. Lastly, the classifier was capable of recognizing highly impactful symptoms like FI, which are frequently unaddressed as patients are often too embarrassed to express them. Thus, these resulting groups captured the ranges of coexisting symptoms while still accounting for the complicated symptomatic overlap of real-world patients, something no other ML categorization system has done thus far.

Our findings here are novel and timely, with the potential to improve diagnosis and treatment and overcome the significant burden these conditions place on patients and the healthcare system. While various approaches to subclassifying urinary symptoms and genitourinary pain exist,^12,22-27^ most require detailed information (patient demographics, physical exam findings, imaging, genetic or biochemical markers, or other diagnostic testing results) unavailable or unfamiliar to most practioners outside of specialized clinical settings. In addition, most of these categorization schema evaluated narrow populations without overlapping symptoms, which ignores a large subset of real-world patients. Here, we generate a diagnostic algorithm based on our novel ML-based phenotypic classification that can be used for treatment assignment of a broad care-seeking population with a wide range of urinary and pelvic complaints. By relying only on patient-reported symptoms, this algorithm can be applied by any type of practitioner in any care delivery setting, including telemedicine.

Providing adequate care for the enormous population of women affected by urinary complaints faces numerous obstacles, both at the health system and individual levels. First is the lack of recognition of these symptoms. Patients, even when highly bothered, may not seek care for urinary complaints due to shame, embarrassment, or a feeling that symptoms are unavoidable. Doctors may not ask about these issues, and even if they do, may not be able to address them due to time constraints or knowledge gaps. The prevalence of urinary symptoms recognized in the primary care setting increases dramatically when validated questionnaires are routinely administered,^38^ stressing the need for better, more pervasive, and perhaps more anonymized screening. As a result of these numerous barriers, however, few women seek or receive care.^39^ The development of a digital resource providing diagnostic assistance has the potential to dramatically improve the quality of life of the population, not only through more accurate diagnostics but by allowing rapid implementation of first-line, low-risk treatments and earlier access to specialty care.

Although our results are promising, our algorithm is based on data from a single center; thus, our findings may not be scalable to the population at large. Further, accurate classification of patients is only of clinical value if the treatment based on these diagnostic groupings correlates with clinical improvement, which will need to be addressed in future prospective studies. Also, the current modeling requires patients to answer nearly 50 questions, which may not be achievable in most clinical scenarios. Finally, our approach to classifying patients may not be generalizable to populations in different clinical scenarios, such as those not presenting for urologic care.

Despite these limitations, this study examined a large number of patients (over 500 in each cohort, totaling over 1,000 subjects) who were consecutively included regardless of referral diagnosis or presenting complaints. This means that our cohort not only included a control group, but is likely representative of the true patient population referred for urologic care. Furthermore, this algorithm relies only on validated questionnaire responses. Thus, this novel LUTS classification algorithm can be utilized to assign diagnoses without the need for either sub-specialist evaluation, to which access can be limited, or physical examination, which can be challenging for patients in underserved areas. Given a growing physician shortage and increasing challenges for patients accessing specialist care, digital technology holds great potential to improve the early recognition, diagnosis, and early treatment of functional urologic conditions. Future prospective work with a larger, multi-institutional cohort is needed to improve these algorithms and to allow accurate diagnosis and treatment assignment based on the machine learning-suggested phenotypes. In addition, reduction of the number of questions to the minimum subset possible is needed to make this feasible to perform in a clinical setting. With refinement, however, this approach to care delivery is capable of increasing both the equity and rapidity of access to effective urologic care.

## Conclusions

A data-driven approach to the phenotyping of lower urinary tract syptoms using unsupervised and supervised machine learning approaches is capable of recognizing both simple and highly complex phenotypic patterns in patients seeking care for urinary complaints. The development in this report of a digital tool trained to classify a broad range of subjects with urologic symptoms that can be administered virtually shows promise to improve the recognition and accurate diagnosis of such disorders.

## Data Availability

All data produced in the present study are available upon reasonable request to the authors.

## Abbreviations

(LUTS): Lower urinary tract urinary symptoms
(OAB): overactive bladder
(IC/BPS): interstitial cystitis/painful bladder syndrome
(ML): machine learning
(POP): pelvic organ prolapse
(fGUPI): female Genitourinary Pain Index
(OABq: Pelvic Floor Distress Inventory
(PFDI-20): Overactive Bladder Questionnaire
(ICSI/ICPI): Interstitial Cystitis Symptom and Problem Indices
(UTI): urinary tract infection
(UMAP): Uniform manifold approximation and projection
(UUI): (pelvic floor disorders [PFD]), urgency urinary incontinence
(MUI): mixed urinary incontinence
(FI/MUI): fecal incontinence/mixed urinary incontinence
(MFS): myofascial frequency syndrome
(POP): pelvic organ prolapse
(MPP): myofascial pelvic pain
(Controls/SUI): controls/stress urinary incontinence
(NUPP): non-urologic pelvic pain
(SUI): stress urinary incontinence
(UUI): urgency urinary incontinence

## REFERENCES

1. Coyne KS, Sexton CC, Thompson CL, et al. The prevalence of lower urinary tract symptoms (LUTS) in the USA, the UK and Sweden: results from the Epidemiology of LUTS (EpiLUTS) study. BJU Int. Aug 2009;104(3):352–60. doi:10.1111/j.1464-410X.2009.08427.x

2. Lee UJ, Feinstein L, Ward JB, et al. Prevalence of Urinary Incontinence among a Nationally Representative Sample of Women, 2005-2016: Findings from the Urologic Diseases in America Project. J Urol. Jun 2021;205(6):1718–1724. doi:10.1097/JU.0000000000001634

3. Coyne KS, Sexton CC, Irwin DE, Kopp ZS, Kelleher CJ, Milsom I. The impact of overactive bladder, incontinence and other lower urinary tract symptoms on quality of life, work productivity, sexuality and emotional well-being in men and women: results from the EPIC study. BJU Int. Jun 2008;101(11):1388–95. doi:10.1111/j.1464-410X.2008.07601.x

4. Coyne KS, Wein A, Nicholson S, Kvasz M, Chen CI, Milsom I. Comorbidities and personal burden of urgency urinary incontinence: a systematic review. Int J Clin Pract. Oct 2013;67(10):1015–33. doi:10.1111/ijcp.12164

5. Coyne KS, Wein A, Nicholson S, Kvasz M, Chen CI, Milsom I. Economic burden of urgency urinary incontinence in the United States: a systematic review. Journal of managed care pharmacy : JMCP. Feb 2014;20(2):130–40. doi:10.18553/jmcp.2014.20.2.130

6. Koch LH. Help-seeking behaviors of women with urinary incontinence: an integrative literature review. J Midwifery Womens Health. Nov-Dec 2006;51(6):e39–44. doi:10.1016/j.jmwh.2006.06.004

7. Berry SH, Bogart LM, Pham C, et al. Development, validation and testing of an epidemiological case definition of interstitial cystitis/painful bladder syndrome. Research Support, N.I.H., Extramural Validation Studies. J Urol. May 2010;183(5):1848–52. doi:10.1016/j.juro.2009.12.103

8. NIH. Meeeting on Measurement of Urinary Symptoms http://www.niddk.nih.gov/news/events-calendar/Pages/meeting-on-measurement-of-urinary-symptoms-momus.aspx

9. Clemens JQ, Erickson DR, Varela NP, Lai HH. Diagnosis and Treatment of Interstitial Cystitis/Bladder Pain Syndrome. J Urol. Jul 2022;208(1):34–42. doi:10.1097/JU.0000000000002756

10. Lightner DJ, Gomelsky A, Souter L, Vasavada SP. Diagnosis and Treatment of Overactive Bladder (Non-Neurogenic) in Adults: AUA/SUFU Guideline Amendment 2019. J Urol. Sep 2019;202(3):558–563. doi:10.1097/JU.0000000000000309

11. Benner JS, Nichol MB, Rovner ES, et al. Patient-reported reasons for discontinuing overactive bladder medication. BJU Int. May 2010;105(9):1276–82. doi:10.1111/j.1464-410X.2009.09036.x

12. Andreev VP, Liu G, Yang CC, et al. Symptom Based Clustering of Women in the LURN Observational Cohort Study. J Urol. Dec 2018;200(6):1323–1331. doi:10.1016/j.juro.2018.06.068

13. Clemens J, Calhoun E, Litwin M, et al. Validation of a modified National Institutes of Health chronic prostatitis symptom index to assess genitourinary pain in both men and women. Urology. 2009;74:983–987.e3.

14. Coyne K, Revicki D, Hunt T, et al. Psychometric validation of an overactive bladder symptom and health-related quality of life questionnaire: The OAB-q.. Qual Life Res. 2002;11:563–574.

15. Barber MD, Walters MD, Bump RC. Short forms of two condition-specific quality-of-life questionnaires for women with pelvic floor disorders (PFDI-20 and PFIQ-7). Am J Obstet Gynecol. Jul 2005;193(1):103–13. doi:10.1016/j.ajog.2004.12.025

16. O’Leary MP, Sant GR, Fowler FJ, Whitmore KE, Spolarich-Kroll J. The interstitial cystitis symptom index and problem index. Urology. 1997;49(5):58–63.

17. Ward JHJ. Hierarchical Grouping to Optimize an Objective Function. Journal of the American Statistical Association. 1963;58(301):236–244. doi:10.1080/01621459.1963.10500845

18. Rousseeuw PJ. Silhouettes: A graphical aid to the interpretation and validation of cluster analysis. Journal of Computational and Applied Mathematics. 1987/11/01/ 1987;20:53–65. doi:https://doi.org/10.1016/0377-0427(87)90125-7

19. Grinsztajn L, Oyallon E, Varoquaux G. Why do tree-based models still outperform deep learning on tabular data? arXiv. arXiv; 2022. doi:10.48550/ARXIV.2207.08815

20. Brodersen KH, Ong CS, Stephan KE, Buhmann JM. The Balanced Accuracy and Its Posterior Distribution. presented at: 2010 20th International Conference on Pattern Recognition; 23–26 Aug. 2010 2010;

21. McInnes L, Healy J, Melville J. UMAP: Uniform Manifold Approximation and Projection for Dimension Reduction. arXiv. arXiv; 2018. doi:10.48550/ARXIV.1802.03426

22. Hall SA, Cinar A, Link CL, et al. Do urological symptoms cluster among women? Results from the Boston Area Community Health Survey. BJU Int. May 2008;101(10):1257–66. doi:10.1111/j.1464-410X.2008.07557.x

23. Andreev VP, Helmuth ME, Liu G, et al. Subtyping of common complex diseases and disorders by integrating heterogeneous data. Identifying clusters among women with lower urinary tract symptoms in the LURN study. PloS one. 2022;17(6):e0268547. doi:10.1371/journal.pone.0268547

24. Coyne KS, Matza LS, Kopp ZS, et al. Examining lower urinary tract symptom constellations using cluster analysis. BJU Int. May 2008;101(10):1267–73. doi:10.1111/j.1464-410X.2008.07598.x

25. Ueda T, Hanno PM, Saito R, Meijlink JM, Yoshimura N. Current Understanding and Future Perspectives of Interstitial Cystitis/Bladder Pain Syndrome. Int Neurourol J. Jun 2021;25(2):99–110. doi:10.5213/inj.2142084.042

26. Lai HH, Jemielita T, Sutcliffe S, et al. Characterization of Whole Body Pain in Urological Chronic Pelvic Pain Syndrome at Baseline: A MAPP Research Network Study. J Urol. Sep 2017;198(3):622–631. doi:10.1016/j.juro.2017.03.132

27. Gross J, Vetter JM, Lai HH. Clustering of patients with overactive bladder syndrome. BMC urology. Mar 19 2021;21(1):41. doi:10.1186/s12894-021-00812-9

28. Mwesigwa PJ, Jackson NJ, Caron AT, et al. Unsupervised Machine Learning Approaches Reveal Distinct Phenotypes of Perceived Bladder Pain: A Pilot Study. Front Pain Res (Lausanne). Nov 2021;2doi:10.3389/fpain.2021.757878

29. Ackerman AL, Lai HH, Parameshwar PS, Eilber KS, Anger JT. Symptomatic Overlap in Overactive Bladder and Interstitial Cystitis/Painful Bladder Syndrome -development of a new algorithm. BJU Int. Sep 25 2018;doi:10.1111/bju.14568

30. Lai HH, Vetter J, Jain S, Gereau RWt, Andriole GL. The overlap and distinction of self-reported symptoms between interstitial cystitis/bladder pain syndrome and overactive bladder: a questionnaire based analysis. J Urol. Dec 2014;192(6):1679–85. doi:10.1016/j.juro.2014.05.102

31. Karjalainen PK, Tolppanen AM, Mattsson NK, Wihersaari OAE, Jalkanen JT, Nieminen K. Pelvic organ prolapse surgery and overactive bladder symptoms-a population-based cohort (FINPOP). Int Urogynecol J. Jan 2022;33(1):95–105. doi:10.1007/s00192-021-04920-w

32. Chow D, Rodriguez LV. Epidemiology and prevalence of pelvic organ prolapse. Current opinion in urology. Jul 2013;23(4):293–8. doi:10.1097/MOU.0b013e3283619ed0

33. Bradley CS, Zimmerman MB, Qi Y, Nygaard IE. Natural history of pelvic organ prolapse in postmenopausal women. Obstet Gynecol. Apr 2007;109(4):848–54. doi:10.1097/01.AOG.0000255977.91296.5d

34. Handa VL, Garrett E, Hendrix S, Gold E, Robbins J. Progression and remission of pelvic organ prolapse: a longitudinal study of menopausal women. Am J Obstet Gynecol. Jan 2004;190(1):27–32. doi:10.1016/j.ajog.2003.07.017

35. Faubion SS, Shuster LT, Bharucha AE. Recognition and management of nonrelaxing pelvic floor dysfunction. Mayo Clinic proceedings. Feb 2012;87(2):187–93. doi:10.1016/j.mayocp.2011.09.004

36. Meister MR, Sutcliffe S, Badu A, Ghetti C, Lowder JL. Pelvic floor myofascial pain severity and pelvic floor disorder symptom bother: is there a correlation? Am J Obstet Gynecol. Sep 2019;221(3):235 e1–235 e15. doi:10.1016/j.ajog.2019.07.020

37. Ackerman AL, Jackson NJ, Caron AT, Kaufman MR, Routh JC, Lowder JL. Myofascial Frequency Syndrome: A novel syndrome of bothersome lower urinary tract symptoms associated with myofascial pelvic floor dysfunction. In Submission. 2023;

38. Schussler-Fiorenza Rose SM, Gangnon RE, Chewning B, Wald A. Increasing Discussion Rates of Incontinence in Primary Care: A Randomized Controlled Trial. J Womens Health (Larchmt). Nov 2015;24(11):940–9. doi:10.1089/jwh.2015.5230

39. Malani P, Kullgren J, Solway E. National Poll on Healthy Aging (NPHA). 2019.

